# Immunogenicity and reactogenicity of SARS-CoV-2 vaccines in people living with HIV: a nationwide prospective cohort study in the Netherlands

**DOI:** 10.1101/2022.03.31.22273221

**Authors:** Kathryn S. Hensley, Marlou J. Jongkees, Daryl Geers, Corine H. GeurtsvanKessel, Yvonne M. Mueller, Virgil A.S.H. Dalm, Grigorios Papageorgiou, Hanka Steggink, Alicja Gorska, Susanne Bogers, Jan G. den Hollander, Wouter F.W. Bierman, Luc B.S. Gelinck, Emile F. Schippers, Heidi S.M. Ammerlaan, Marc van der Valk, Marit G.A. van Vonderen, Corine E. Delsing, Elisabeth H. Gisolf, Anke H.W. Bruns, Fanny N. Lauw, Marvin A.H. Berrevoets, Kim C.E. Sigaloff, Robert Soetekouw, Judith Branger, Quirijn de Mast, Adriana J.J. Lammers, Selwyn H. Lowe, Rory D. de Vries, Peter D. Katsikis, Bart J.A. Rijnders, Kees Brinkman, Anna H.E. Roukens, Casper Rokx

## Abstract

**Background:** Vaccines can be less immunogenic in people living with HIV (PLWH), but for SARS-CoV-2 vaccinations this is unknown.

**Methods and Findings:** A prospective cohort study to examine the immunogenicity of BNT162b2, mRNA-1273, ChAdOx1-S and Ad26.COV2.S vaccines in adult PLWH, without prior COVID-19, compared to HIV-negative controls. The primary endpoint was the anti-spike SARS-CoV-2 IgG response after mRNA vaccination. Secondary endpoints included the serological response after vector vaccination, anti-SARS-CoV-2 T-cell response and reactogenicity.

Between February-September 2021, 1154 PLWH (median age 53 [IQR 44-60], 86% male) and 440 controls (median age 43 [IQR 33-53], 29% male) were included. 884 PLWH received BNT162b2, 100 mRNA-1273, 150 ChAdOx1-S, and 20 Ad26.COV2.S. 99% were on antiretroviral therapy, 98% virally suppressed, and the median CD4+T-cell count was 710 cells/µL [IQR 520-913]. 247 controls received mRNA-1273, 94 BNT162b2, 26 ChAdOx1-S and 73 Ad26.COV2.S. After mRNA vaccination, geometric mean concentration was 1418 BAU/mL in PLWH (95%CI 1322-1523), and after adjustment for age, sex, and vaccine type, HIV-status remained associated with a decreased response (0.607, 95%CI 0.508-0.725). In PLWH vaccinated with mRNA-based vaccines, higher antibody responses were predicted by CD4+T-cell counts 250-500 cells/µL (2.845, 95%CI 1.876-4.314) or >500 cells/µL (2.936, 95%CI 1.961-4.394), whilst a viral load >50 copies/mL was associated with a reduced response (0.454, 95%CI 0.286-0.720). Increased IFN-γ, CD4+, and CD8+T-cell responses were observed after stimulation with SARS-CoV-2 spike peptides in ELISpot and activation induced marker assays, comparable to controls. Reactogenicity was generally mild without vaccine-related SAE.

**Conclusion:** After vaccination with BNT162b2 or mRNA-1273, anti-spike SARS-CoV-2 antibody levels were reduced in PLWH. To reach and maintain the same serological responses and vaccine efficacy as HIV-negative controls, additional vaccinations are probably required.

## Introduction

At the end of 2019, severe acute respiratory syndrome coronavirus 2 (SARS-CoV-2) emerged and the ensuing and ongoing pandemic led to the loss of millions of lives. Highly effective vaccines were quickly implemented, and mass vaccination campaigns have become the cornerstone to prevent fatal Coronavirus Disease (COVID-19) to quell this pandemic, four of which are currently approved for use in the Netherlands [1-5].

After the SARS-CoV-2 vaccine registration trials, vaccinations were rolled out globally. However, people with immune deficiencies were only sporadically included in the original phase three SARS-CoV-2 vaccination trials. Subsequent reports indicated markedly diminished responses in solid organ transplant recipients, and lower responses in haemodialysis patients, stem cell recipients or patient groups on specific immunosuppressant drugs for immune disorders [6-9].

HIV infection is associated with worse COVID-19 outcomes although the underlying mechanism is not yet clear [10]. In most countries, people living with HIV (PLWH) were therefore prioritised for SARS-CoV-2 vaccination. PLWH show diminished responses to a wide variety of vaccines such as hepatitis B, and seasonal influenza vaccines compared to HIV-uninfected individuals [11, 12]. This might also hold true for SARS-CoV-2 vaccines. Indicative of potential lower responses to SARS-CoV-2 vaccines could be that after SARS-CoV-2 infection, lower IgG concentrations and neutralising antibody titres were found in PLWH compared to controls [13]. Data are scarce on SARS-CoV-2 vaccination responses in PLWH. Small studies using the ChAdOx1-S vaccine in the UK and South Africa in relatively young PLWH with high CD4+ T-cell counts gave comparable responses to controls [14, 15]. As for BNT162b2, similar results were shown in a limited number of PLWH [16-18]. The identification of risk factors for a reduced response to SARS-CoV-2 vaccines in PLWH is important as it will help to improve vaccination strategies in PLWH. A good understanding of vaccination response in PLWH becomes even more important now that variants of concern (VOC) continue to arise and partially escape vaccine induced immunity [19]. In this study we investigated the immunogenicity and reactogenicity of SARS-CoV-2 vaccinations in PLWH with the vaccines currently approved in the Netherlands; BNT162b2, mRNA-1273, ChAdOx1-S, or Ad26.COV2.S.

## Methods

### Study design and participants

We performed a prospective observational cohort study in 22 of the 24 HIV treatment centres in the Netherlands. Participants were recruited via treating physicians or nurses specialised in HIV-care. To be eligible for participation, a subject had to be 18 years or older, had a confirmed HIV infection and was invited for SARS-CoV-2 vaccination by Dutch public health services. Participants with a history of previous SARS-CoV-2 infection demonstrated by PCR or detectable SARS-CoV-2 antibodies in serum before vaccination were excluded. Inclusion was stratified and monitored to best represent represents the Dutch population of PLWH (S1 appendix) [20].

Participants received either BNT162b2, mRNA-1273, ChAdOx1-S, or Ad26.COV2.S according to manufacturer’s regulations as part of the Dutch SARS-CoV-2 vaccination campaign (S1 appendix). Vaccination response data from HIV-negative controls were obtained from two separate concurrent studies (n=440; S1 appendix) [21].

### Clinical procedures

Blood samples were collected up to six weeks before vaccination (T0) and four to six weeks after the completed vaccination schedule (T2). In a subgroup, additional blood samples were collected, 21 days (+/- three days) after the first vaccination for serology (T1) or peripheral blood mononuclear cells (PBMCs) at any of the study visits. Participants are scheduled for longitudinal blood sampling for additional analyses during two years which will be reported separately.

Study variables were collected in an electronic case record file (S1 appendix). Participants received a paper diary or a link to an online questionnaire to enter adverse events (AE) from a predefined list and medication use for AE the seven days following each vaccination.

### Laboratory procedures

All serum samples were collected via venepuncture at participating centres. Serum samples before vaccination were analysed at the laboratory of the individual treating centres with the available SARS-CoV-2 antibody test (S1 appendix). Serum samples post-vaccination were transported for testing at the department of Viroscience, Erasmus Medical Centre, the Netherlands. The presence of binding antibodies against the SARS-CoV-2 Spike (S1) were quantified with a validated IgG Trimeric chemiluminescence immunoassay (DiaSorin Liaison) with a lower limit of detection at 4.81 binding antibody units per millilitre (BAU/mL).

PBMCs were isolated by density gradient centrifugation (Ficoll-Hypaque, GE Healthcare life sciences) and collected in RPMI-1640 (Life Technologies) supplemented with 3% foetal bovine serum (FBS). PBMCs were washed three times, frozen in freezing media (90% FBS, 10% dimethyl sulfoxide (DMSO)) and stored in liquid nitrogen until use.

We used enzyme linked immune absorbent spots (ELISpot) to quantify interferon (IFN)-γ secretion in response to SARS-CoV-2 peptides. ELISpot assays were performed on cryopreserved PBMCs using a commercial double colour kit (ImmunoSpot®, Cellular Technology Limited) including peptide pools containing spike and nucleocapsid protein (S1 appendix). Results are expressed as spot forming cells (SFCs) per million PBMCs, calculated by subtracting mean response after MOG stimulation.

T-cell responses were further characterised by activation induced marker (AIM) assay. PBMCs were incubated with SARS-CoV-2 peptide pools covering the entire S-protein of the WuhanHu1 (wild-type) or B.1.617.2 (delta) variant. Following stimulation, cells were stained and measured by flow cytometry (FACSLyric, BD; S1 appendix). SARS-CoV-2-reactive T-cells were identified as CD137+OX40+ for CD4+ subtype or CD137+CD69+ for CD8+ subtype. On average, 300,000 cells were measured. The gating strategy can be found in S1 Fig.

### Outcomes

The primary outcome was the height of the anti-spike SARS-CoV-2 IgG response in PLWH 4-6 weeks after the completed vaccination schedule with BNT162b2 or mRNA-1273. This endpoint was chosen because primarily mRNA vaccines were allocated to PLWH in the Netherlands. Secondary outcomes included the antibody response in PLWH after the completed vaccination schedule with ChAdOx1-S and Ad26.COV2.S, and variables associated with the height of antibody level (S1 appendix). Variables associated with hyporesponse and non-response were also analysed (S1 appendix). We defined a hyporesponse as 50-300 BAU/mL and non-response as <50 BAU/mL, based on previous studies which showed correlation of antibody concentration of 300 BAU/mL with a neutralising capacity of 1:40.[9, 22] In the subgroup, anti-spike SARS-CoV-2 specific T-cell response, and antibody response 21 days after the first vaccine dose were evaluated. Lastly, we evaluated the tolerability of the vaccines by monitoring vaccine-related AE (S1 appendix).

### Sample size and statistical analysis plan

When the study started, we did not have a confirmed availability of a control group due to the rapid initiation of the national immunisation campaign. We justified the sample size by calculating that 556 PLWH receiving mRNA vaccines would be sufficient to detect a serological response rate of 90% or lower compared to a hypothetical 95% response rate in controls with >80% power. When the control group was confirmed, and before the data lock and endpoint analyses, we amended the protocol to update the sample size calculation. Accounting for the imbalance in the number of controls versus PLWH with BNT162b2 and mRNA-1273 vaccinations, we found that 286 controls were sufficient to detect a 20% lower antibody response in PLWH with >80% power and alpha 5%.

Median (interquartile range (IQR)), or n (%) for descriptive data were used. A multivariable linear regression model was used for the analysis of the anti-spike SARS-CoV-2 IgG. The outcome was transformed using the natural logarithm plus one unit: ln(anti-spike SARS-CoV-2 IgG+1) in order to meet the model assumptions. All patients of which the sample was received in the central laboratory for testing who completed the vaccination scheme were included in the analysis (per protocol). The difference between PLWH and controls was captured by the corresponding regression coefficient and 95%CI. The model was further adjusted for differences in vaccine type, age, and sex. A multivariable linear regression model was also used to quantify the difference in ln(anti-spike SARS-CoV-2 IgG+1) between PLWH versus controls for the subset in the sample vaccinated with vector vaccines. A similar model was used to quantify the effect of age, sex, vaccine type, most recent CD4+ T-cell count, and antibody concentration in PLWH. In addition, multivariable logistic regression models were used to calculate odds ratios with 95%CI for the effects of sex, age, nadir CD4+ T-cell count, most recent CD4+ T-cell count, HIV-RNA viral load, and vaccine group in PLWH on having a hyporesponse or a non-response. In subgroup participants we evaluated differences from baseline to T1 and T1 to T2, as well as AIM data comparing T0 and T2 time points, by Wilcoxon matched-pairs signed rank test. ELISpot T0 to T2 data and AIM data comparing PLWH and controls were analysed by Mann-Whitney-U tests.

Data were analysed using IBM SPSS Statistics 25, R (v. 4.1.2), and GraphPad Prism 8. Flow cytometry data were analysed using FlowJo software version 10.8.1.

### Ethical considerations

The trial was performed in accordance with the principles of the Declaration of Helsinki, Good Clinical Practice guidelines, and in accordance with the Dutch Medical Research Involving Human Subjects Act (WMO). Written informed consent was obtained from all participants. The trial was reviewed and approved by the Medical research Ethics Committees United Nieuwegein (MEC-U, reference 20.125). The trial was registered in the Netherlands Trial Register (NL9214).

## Results

### Baseline characteristics

Between 14 February and 7 September 2021, 1269 PLWH were enrolled (Fig 1.). At sampling before vaccination, 53 (4.2%) PLWH had antibodies against SARS-CoV-2 above test cut-off and were excluded. Overall, 30 PLWH were lost to follow-up (2.4%), five decided against vaccination after inclusion (0.4%), eight withdrew from the study (0.6%), and ten samples were not stored adequately or not received in the central laboratory for testing (0.8%). One participant was excluded from the final analysis because they received two different vaccines (ChAdox1-S and BNT162b2). In the final analysis, 76.6% PLWH received BNT162b2, 8.7% mRNA-1273, 13.0% ChAdOx1-S and 1.7% Ad26.COV2.S. Included PLWH had a median age of 53 [IQR 44-60], 85.5% were men and had a median CD4+ T-cell count before vaccination of 710 cells/µL [IQR 520-913] (Table 1). The vast majority (99.0%) was on cART and had a suppressed plasma HIV-RNA (97.7% <50 copies/mL). The control group consisted of 440 people of which 94 were vaccinated with BNT162b2 (21.4%), 247 with mRNA-1273 (56.1%), 26 ChAdOx1-S (5.9%) and 73 Ad26.COV2.S (16.6%). Their median age was 43 [IQR 33-53] and 28.6% were men. The age distribution across groups differed, with the majority of PLWH receiving ChAdOx1-S being 55-65 years of age compared to 15-25% for the other vaccines (S1 Table). Age differences were also seen between groups, with less participants of older ages in the control group. Between the control group and PLWH, there was also a difference in inclusion of sex. All other factors were similar across groups. In the subgroup of PLWH, baseline characteristics reflected the characteristics of included PLWH, with most participants receiving the BNT162b2 vaccine (66.0-95.3%) and being male (76.6-85.7%) (S2 Table).

**Table 1:** Baseline characteristics of HIV negative controls and PLWH.

|  | HIV negative participants |  |  | People living with HIV |  |  |
| --- | --- | --- | --- | --- | --- | --- |
|  | Overall<br>N = 440 | mRNA<br>vaccines<sup>1</sup><br>N = 341<br>(77.5%) | Vector<br>vaccines<sup>2</sup><br>N = 99<br>(22.5%) | Overall<br>N = 1154 | mRNA<br>vaccines<sup>1</sup><br>N = 984<br>(85.3%) | Vector<br>vaccines<sup>2</sup><br>N = 170<br>(14.7%) |
| **Sex assigned at birth** |  |  |  |  |  |  |
| Male | 126 (28.6%) | 104 (30.5%) | 22 (22.2%) | 987 (85.5%) | 839 (85.2%) | 148 (87.1%) |
| Female | 314 (71.4%) | 237 (69.5%) | 77 (77.8%) | 167 (14.5%) | 145 (14.8%) | 22 (12.9%) |
| **Age category** |  |  |  |  |  |  |
| 18-55 years | 352 (80.0%) | 284 (83.3%) | 68 (68.7%) | 703 (60.9%) | 674 (68.5%) | 29 (17.1%) |
| 56-65 years | 74 (16.8) | 43 (12.6%) | 31 (31.3%) | 291 (25.2%) | 157 (16.0%) | 134 (79.3%) |
| 65+ years | 14 (3.2%) | 14 (4.1%) | 0 | 160 (13.9%) | 153 (15.6%) | 7 (4.1%) |
| **On cART** |  |  |  |  |  |  |
| Yes | NA | NA | NA | 1142 (99.0%) | 972 (98.8%) | 170 (100%) |
| No | NA | NA | NA | 12 (1.0%) | 12 (1.2%) | 0 |
| **Most recent plasma HIV viral load** |  |  |  |  |  |  |
| <50 copies/mL | NA | NA | NA | 1127 (97.7%) | 960 (97.6%) | 167 (98.2%) |
| ≥50 copies/mL | NA | NA | NA | 27 (2.3%) | 24 (2.4%) | 3 (1.8%) |
| **Most recent CD4+ T-cell count** |  |  |  |  |  |  |
| <250 cells/μL | NA | NA | NA | 41 (3.6%) | 35 (3.6%) | 6 (3.5%) |
| 250-500 cells/μL | NA | NA | NA | 224 (19.4%) | 189 (19.2%) | 35 (20.6%) |
| >500 cells/μL | NA | NA | NA | 889 (77.0%) | 760 (77.2%) | 129 (75.9%) |
| **Nadir CD4+ T-cell count** |  |  |  |  |  |  |
| <250 cells/μL | NA | NA | NA | 443 (38.4%) | 365 (37.1%) | 78 (45.9%) |
| 250-500 cells/μL | NA | NA | NA | 376 (32.6%) | 330 (33.5%) | 46 (27.1%) |
| >500 cells/μL | NA | NA | NA | 152 (13.2%) | 133 (13.5%) | 19 (11.2%) |
| Unknown | NA | NA | NA | 183 (15.9%) | 156 (15.9%) | 27 (15.9%) |
| **Days between doses\*** | 28 (25-28) | 28 (25-28) | 56 (56-70) | 35 (35-36) | 35 (35-36) | 70 (49-77) |

Data are n (%) or median (IQR). 1BNT162b2, mRNA-1273. 2ChAdOx1-S, Ad26.COV2.S \*Does not apply
for Ad26.COV2.S. NA: not applicable, PLWH: people living with HIV, IQR: interquartile range, cART:
combination antiretroviral therapy

**Fig 1.**
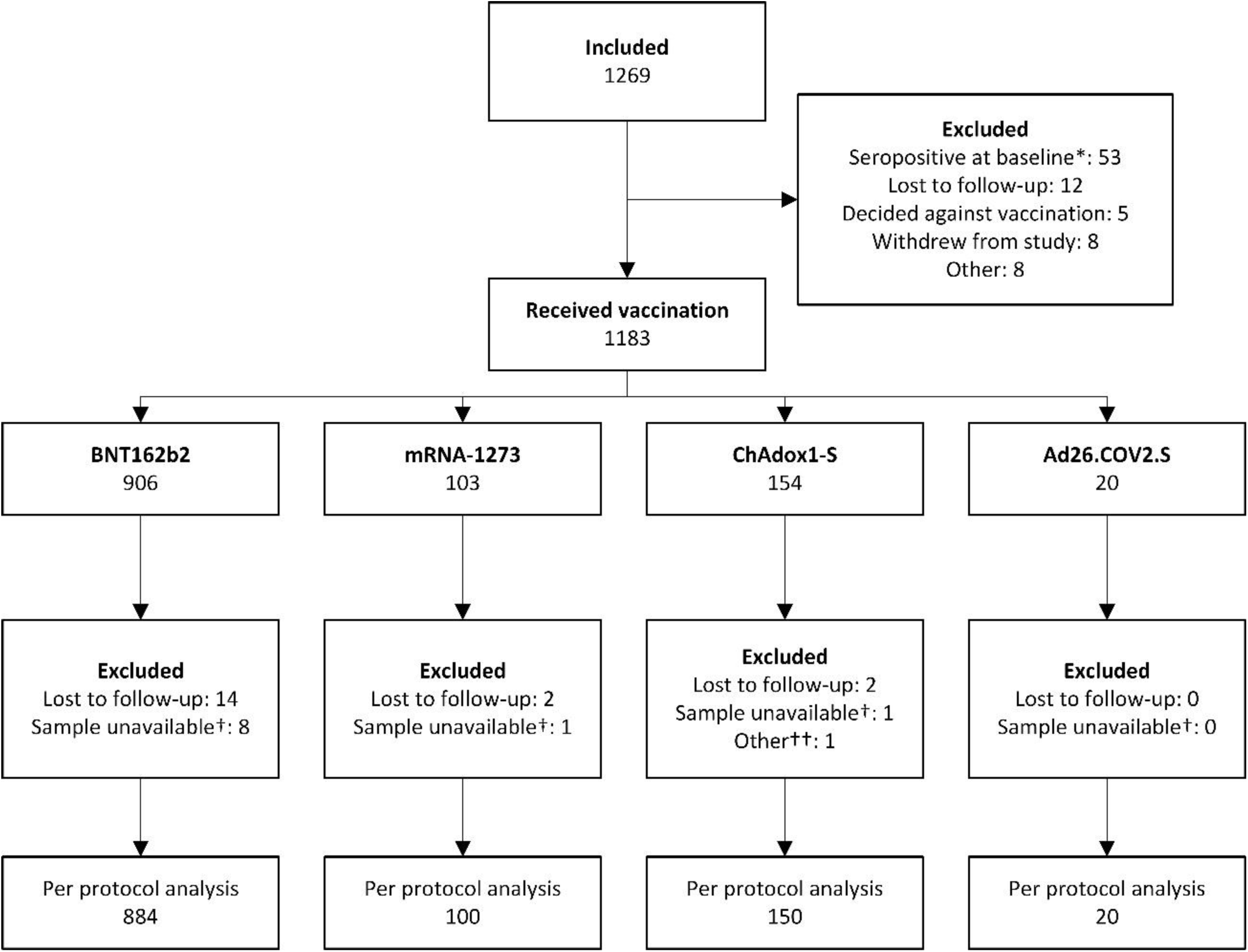
Flow chart of inclusions of PLWH. *Participants that tested positive for SARS-CoV-2 antibodies in serum at baseline measurement were excluded from further participation †Samples were not stored adequately or sent to be analysed in the central laboratory at the Erasmus MC †† One participant received a combination ChAdOx1-S and BNT162b2 vaccines and was therefore not included in the per protocol analysis. PLWH: people living with HIV

### Humoral responses

In all vaccines investigated, antibody concentrations were lower in PLWH compared to controls (Fig 2). In participants vaccinated with an mRNA vaccine, the geometric mean concentration (GMC) was 1418 BAU/mL in PLWH (95%CI 1322-1523) and 3560 BAU/mL in controls (95%CI 3301-3840). With regard to the primary endpoint, after adjusting for age, vaccine, and sex, HIV infection remained associated with a 39.35% lower antibody concentration in PLWH compared to controls receiving an mRNA vaccine (0.607, 95%CI 0.508-0.725). The estimated effect of having an HIV infection was larger than the estimated effects of male sex (23.05% (0.769 95%CI 0.667-0.888)) or age >65 years (35.47% (0.645 95%CI 0.544-0.765)), which were all significantly associated with a worse vaccine response (p<0.001 for all).

**Fig 2:**
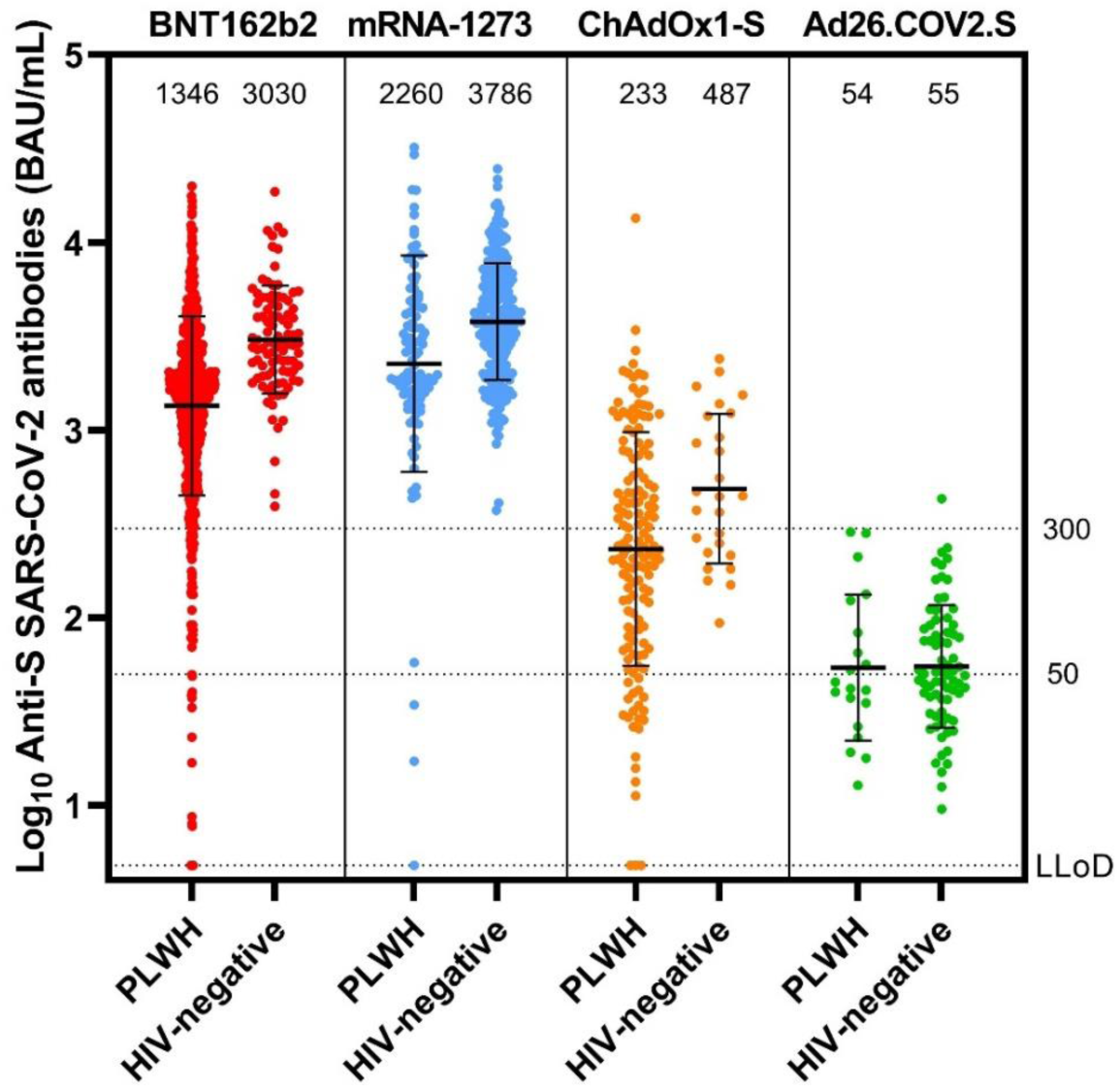
Antibody concentration in PLWH and controls after vaccination. Anti-spike SARS-CoV-2 binding antibodies after BNT162b2, mRNA-1273, ChAdOx1-S or Ads26.COV2.S vaccination in PLWH and HIV-negative control group. The horizontal bar shows the geometric mean concentration, also indicated above the graphs, with error bars showing geometric standard deviation. The dotted lines show the lower limit of detection of the performed test (4.81 BAU/mL), non-response cut-off (50 BAU/mL) and hyporesponse cut-off (300 BAU/mL). S: spike, BAU/mL: binding antibodies per millilitre, PLWH: people living with HIV, LLoD: lower limit of detection

Regarding factors possibly related to antibody responses in PLWH who received mRNA vaccines, receiving mRNA-1273 was associated with a 57.15% higher response (1.572, 95%CI 1.225-2.018, p<0.001). Male sex (0.693 95%CI 0.555-0.865, p=0.001) and age over 65 years (0.654 95%CI 0.526-0.814, p<0.001) were associated with lower responses of 30.72%, and 34.56% respectively. We found no association between nadir CD4+ T-cell count and antibody responses, but found a significant effect of 54.62% lower antibodies when the HIV-RNA level was over 50 copies/mL (0.454, 95%CI 0.286-0.720, p=0.001). The largest effect on antibody levels was associated with current CD4+ T-cell counts between 250 and 500 cells/µL (2.845 95%CI 1.876-4.314) and over 500 cells/µL (2.936 95%CI 1.961-4.394) (both p<0.001) with increased antibody concentrations of 184.34% and 193.59% respectively.

In participants receiving vector vaccines, after adjustment, HIV was significantly associated with a 39.47% lower antibody response, comparable to that in mRNA vaccines (0.605 95%CI 0.387-0.945, p=0.027). Within this group of PLWH receiving vector vaccines, unlike mRNA vaccines, age, sex, and a detectable viral load were not associated with antibody responses, but receiving Ad26.COV2.S was associated with a 87.67% lower response (0.123 95%CI 0.051-0.300, p<0.001) and a recent CD4+ T-cell count of 250 to 500 or over 500 cells/µL were associated with better responses (both p<0.001).

Hyporesponse and nonresponse percentages were lower for every vaccine group in PLWH compared to controls (S1 Table). In PLWH, after adjustment, receiving a vector vaccine (p<0.001, OR 0.036), age over 65 (p<0.001, OR 0.282) and a viral load over 50 copies/mL (p=0.017, OR 0.266) were associated with an antibody response under 300 BAU/mL. Most recent CD4+ T-cell count between 250 and 500 cells/µL or over 500 cells/µL were significantly associated with an antibody response of 300 BAU/mL or higher (both p<0.001 OR 8.143 and 9.177 respectively). Sex and nadir CD4+ T-cell count were not associated. When looking at non-response in PLWH, receiving a vector vaccine (p<0.001, OR 0.034) was associated with an antibody response <50 BAU/mL. Being 56 to 65 years of age (p=0.025, OR 2.919) and most recent CD4+ T-cell count between 250 and 500 cells/µL or over 500 cells/µL (both p<0.001 OR 7.810 and 15.853 respectively) were significantly associated with an antibody response of more than 50 BAU/mL. Sex, viral load and nadir CD4+ T-cell count were not associated.

In the subgroup of PLWH in whom extra sampling was performed 21 days after the first vaccination (n=43, 95.3% with BNT162b2), we found response rates of 83.3% with a GMC of 148 BAU/ml at T1 and 1952 BAU/mL at T2 (S2 Fig).

### Cellular responses

In the ELISpot assay, spike-specific T-cell responses measured as IFN-γ, after deduction of MOG, increased from a median of 27.5 SFCs per million PBMCs before vaccination to 152.5 SFC post-vaccination (p=0.0023) (Fig 3A). In PLWH, stimulating PBMC with the negative control peptide pool MOG already induced a IFN-γ-response, resulting in high background (S3 Fig). No responses in nucleocapsid were seen after subtraction of MOG. Additionally, CD4+ and CD8+ T-cell responses were assessed in an AIM assay. SARS-CoV-2 specific CD4+ T-cell (CD137+OX40+) and CD8+ T-cell responses (CD137+CD69+) both increased compared to baseline following vaccination and after correction for DMSO (p=0.0049 and p=0.0078 respectively) (Fig 3C and 3D). CD4+ and CD8+ T cell responses in PLWH against the delta variant were similar before and after vaccination. Importantly, both CD4+ and CD8+ T-cell responses were of similar magnitude between PLWH and HIV negative individuals.

**Fig 3:**
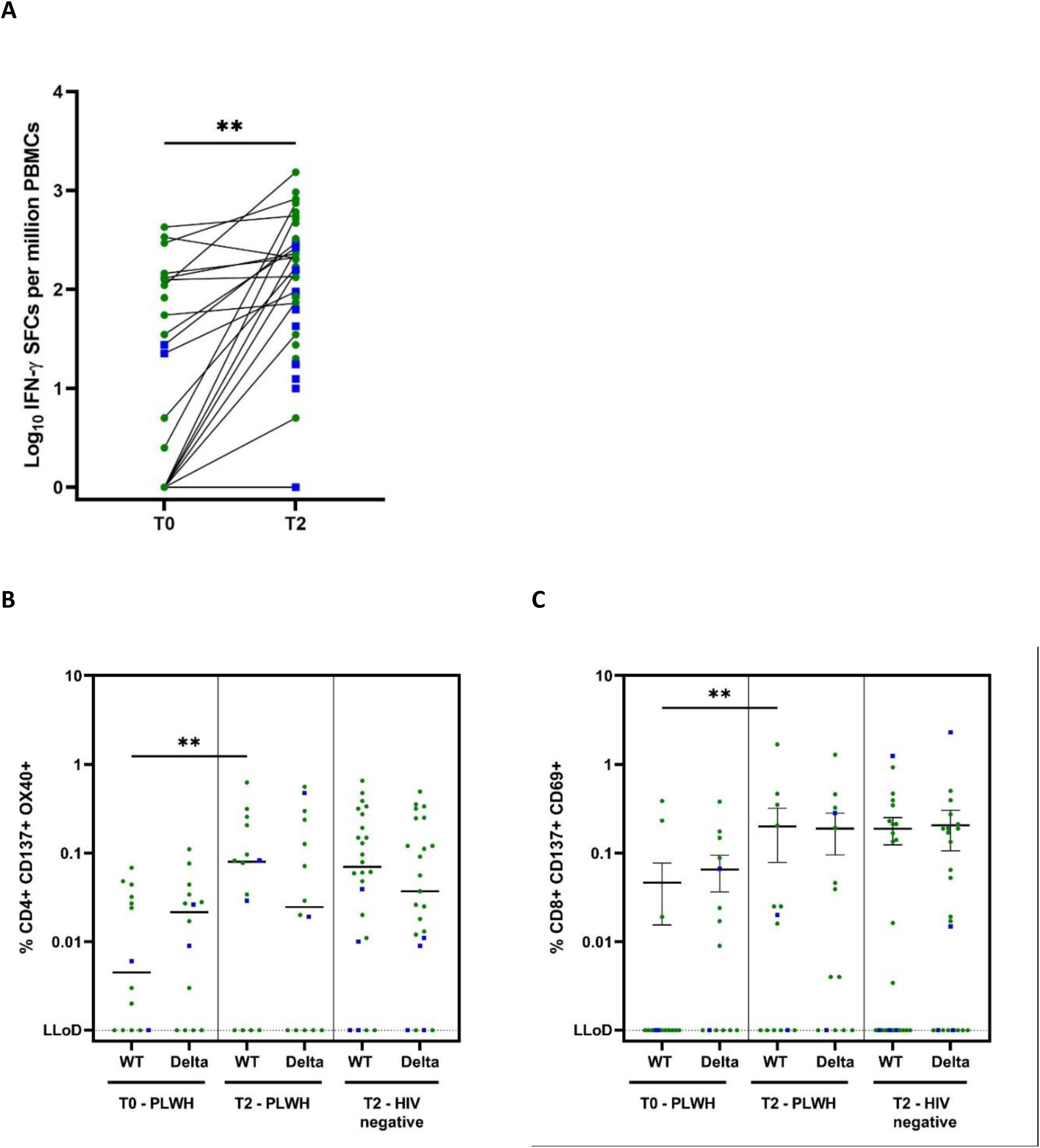
Cellular immune responses against SARS-CoV-2 in subgroup participants (PLWH) **A** Cellular immune response to wild type spike by ELISpot assay (T0 n=23, T2 n=45), IFN-γ SFC after subtraction of MOG. Statistics performed using Mann Whitney test p=0.0023 **B** Cellular immune response to wild type and delta spike in AIM assay (n=14), percentage of CD4+ CD137+ OX40+ T-cells after subtraction of DMSO. Dotted line shows the LLoD at 0.001%. Statistics between T0 and T2 WT in PLWH performed using Wilcoxon matched-pairs sign rank test (p=0.0049), T0 and T2 delta: ns. Statistics between PLWH and controls with Mann Whitney test: ns. **C** Cellular immune response to wild type and delta spike in AIM assay (n=14), percentage of CD8+ CD137+ CD69+ T-cells after subtraction of DMSO. Dotted line shows the LLoD at 0.001%. Statistics performed using Wilcoxon matched-pairs sign rank test (p=0.0078), T0 and T2 delta: ns. Statistics between PLWH and controls with Mann Whitney test: ns. Green circles: mRNA vaccines, Blue squares: vector based vaccines. T0: before vaccination, T2: 4-6 weeks after second vaccination, PLWH: people living with HIV, IFN: interferon, SFC: spot forming cells, PBMCs: peripheral blood mononuclear cells, MOG: myelin-oligodendrocyte glycoprotein, ELISPot: enzyme-linked immune absorbent spot, AIM: activation induced marker, DMSO: dimethyl sulfoxide, WT: wildtype, ns: not significant LLoD: lower limit of detection

### Reactogenicity

In PLWH, the questionnaire was completed 1039 (90.0%) times after the first vaccination and 1026 (90.4%) after the second vaccination. Overall, more than half (52.4%) of the participants reported any AE (Table 2). For those who received two dosages, the frequency of AE did not increase after the second vaccination (from 55.2% to 49.6% respectively). AE after first and second vaccination were for BNT162b2 55.2% and 48.0%, mRNA-1273 62.2% and 70.1%, ChAdOx1-S 51.8% and 46.0%, and after the single Ad26.COV2.S vaccination 47.4%. The most commonly reported local reaction was pain at the injection site (44.4%). The most common systemic reactions were myalgia (13.0%) and headache (18.3%). When AE occurred, most were mild 1159 (65.2%) or moderate 523 (29.4%) in severity and self-limiting (Fig 4). Analgesic or antipyretic drug use was necessary in 346 (16.8%) of all participants, for a cumulative 769 AE, of which 400 (52.0%) were moderate and 57 (7.4%) severe AE. Paracetamol (81.2%) was most commonly used. Ten SAE were reported and all considered unrelated. One participant visited the emergency department three days after vaccination with pain in the arm and chest and shortness of breath in whom a pulmonary embolism was excluded and symptoms resolved. Two participants were admitted for a chronic obstructive pulmonary disease exacerbation. The seven other SAE were elective heart surgery, intestinal perforation after infection, *Campylobacter jejuni* infection, bicycle accident, pyloric stenosis, a hip fracture after a fall and one person died due to a cardiac arrest. There was no discontinuation of the vaccination series due to vaccine related AE.

**Table 2:**
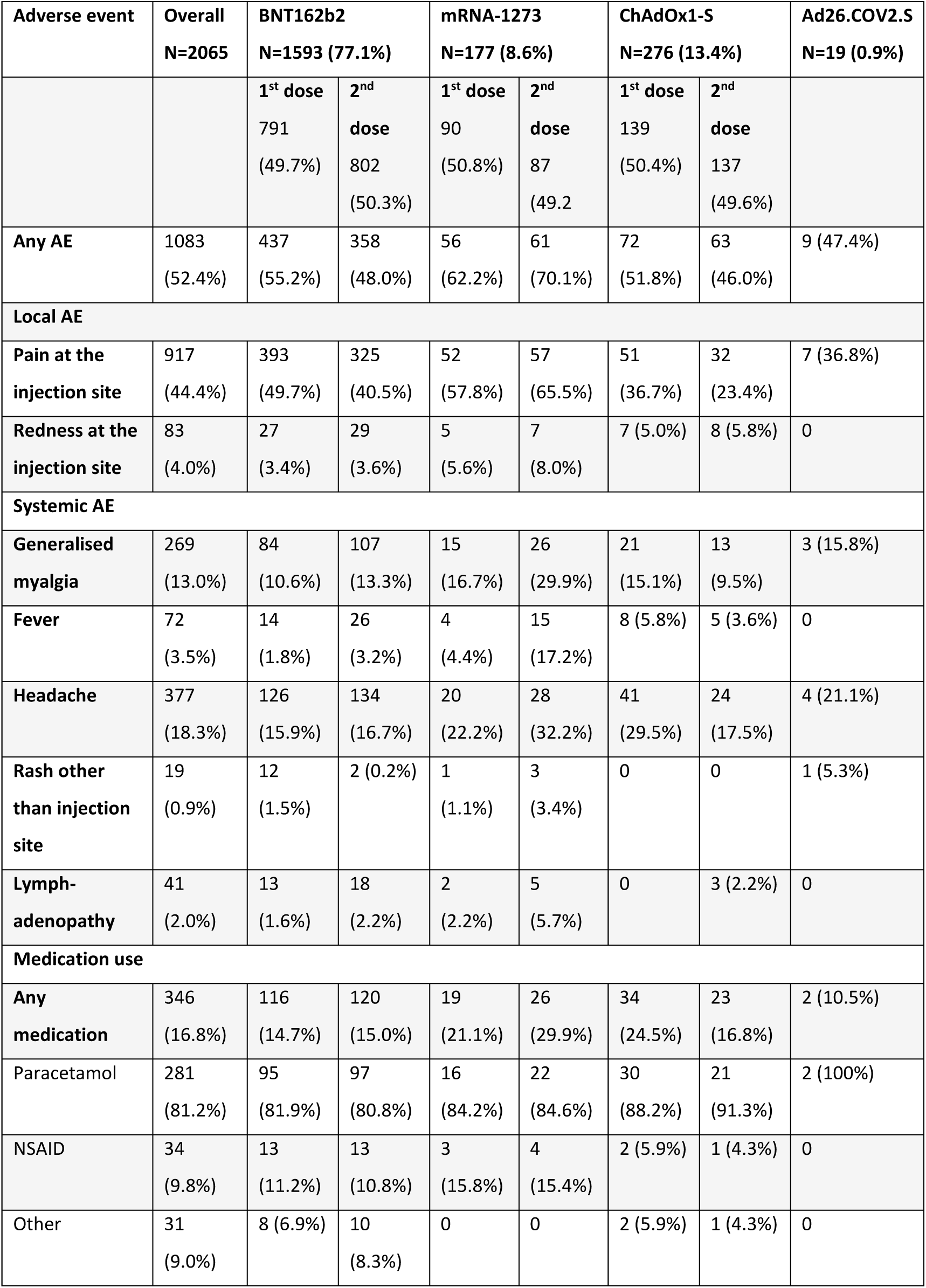
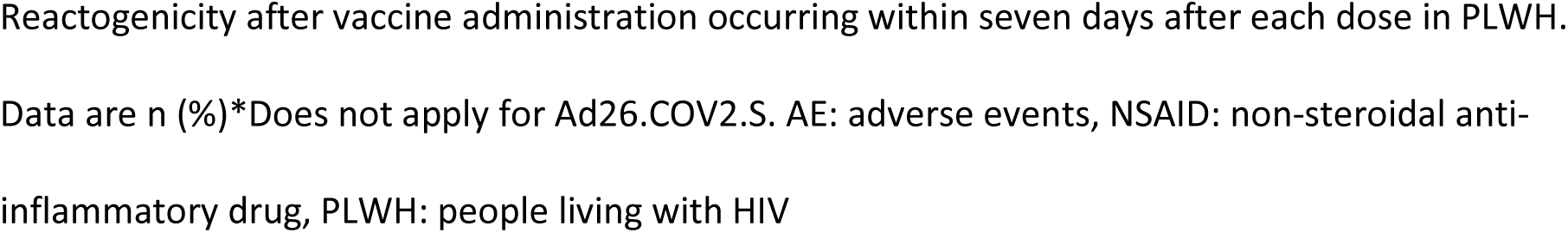
Reactogenicity in PLWH.

| Adverse event | Overall<br>N=2065 | BNT162b2<br>N=1593 (77.1%) |  | mRNA-1273<br>N=177 (8.6%) |  | ChAdOx1-S<br>N=276 (13.4%) |  | Ad26.COV2.S<br>N=19 (0.9%) |
| --- | --- | --- | --- | --- | --- | --- | --- | --- |
|  |  | 1<sup>st</sup> dose<br>791<br>(49.7%) | 2<sup>nd</sup> dose<br>802<br>(50.3%) | 1<sup>st</sup> dose<br>90<br>(50.8%) | 2<sup>nd</sup> dose<br>87<br>(49.2%) | 1<sup>st</sup> dose<br>139<br>(50.4%) | 2<sup>nd</sup> dose<br>137<br>(49.6%) |  |
| **Any AE** | 1083<br>(52.4%) | 437<br>(55.2%) | 358<br>(48.0%) | 56<br>(62.2%) | 61<br>(70.1%) | 72<br>(51.8%) | 63<br>(46.0%) | 9 (47.4%) |
| **Local AE** |  |  |  |  |  |  |  |  |
| **Pain at the injection site** | 917<br>(44.4%) | 393<br>(49.7%) | 325<br>(40.5%) | 52<br>(57.8%) | 57<br>(65.5%) | 51<br>(36.7%) | 32<br>(23.4%) | 7 (36.8%) |
| **Redness at the injection site** | 83<br>(4.0%) | 27<br>(3.4%) | 29<br>(3.6%) | 5<br>(5.6%) | 7<br>(8.0%) | 7 (5.0%) | 8 (5.8%) | 0 |
| **Systemic AE** |  |  |  |  |  |  |  |  |
| **Generalised myalgia** | 269<br>(13.0%) | 84<br>(10.6%) | 107<br>(13.3%) | 15<br>(16.7%) | 26<br>(29.9%) | 21<br>(15.1%) | 13<br>(9.5%) | 3 (15.8%) |
| **Fever** | 72<br>(3.5%) | 14<br>(1.8%) | 26<br>(3.2%) | 4<br>(4.4%) | 15<br>(17.2%) | 8 (5.8%) | 5 (3.6%) | 0 |
| **Headache** | 377<br>(18.3%) | 126<br>(15.9%) | 134<br>(16.7%) | 20<br>(22.2%) | 28<br>(32.2%) | 41<br>(29.5%) | 24<br>(17.5%) | 4 (21.1%) |
| **Rash other than injection site** | 19<br>(0.9%) | 12<br>(1.5%) | 2 (0.2%) | 1<br>(1.1%) | 3<br>(3.4%) | 0 | 0 | 1 (5.3%) |
| **Lymph-adenopathy** | 41<br>(2.0%) | 13<br>(1.6%) | 18<br>(2.2%) | 2<br>(2.2%) | 5<br>(5.7%) | 0 | 3 (2.2%) | 0 |
| **Medication use** |  |  |  |  |  |  |  |  |
| **Any medication** | 346<br>(16.8%) | 116<br>(14.7%) | 120<br>(15.0%) | 19<br>(21.1%) | 26<br>(29.9%) | 34<br>(24.5%) | 23<br>(16.8%) | 2 (10.5%) |
| Paracetamol | 281<br>(81.2%) | 95<br>(81.9%) | 97<br>(80.8%) | 16<br>(84.2%) | 22<br>(84.6%) | 30<br>(88.2%) | 21<br>(91.3%) | 2 (100%) |
| NSAID | 34<br>(9.8%) | 13<br>(11.2%) | 13<br>(10.8%) | 3<br>(15.8%) | 4<br>(15.4%) | 2 (5.9%) | 1 (4.3%) | 0 |
| Other | 31<br>(9.0%) | 8 (6.9%) | 10<br>(8.3%) | 0 | 0 | 2 (5.9%) | 1 (4.3%) | 0 |

Reactogenicity after vaccine administration occurring within seven days after each dose in PLWH. Data are n (%) \*Does not apply for Ad26.COV2.S. AE: adverse events, NSAID: non-steroidal anti-inflammatory drug, PLWH: people living with HIV

**Fig 4:**
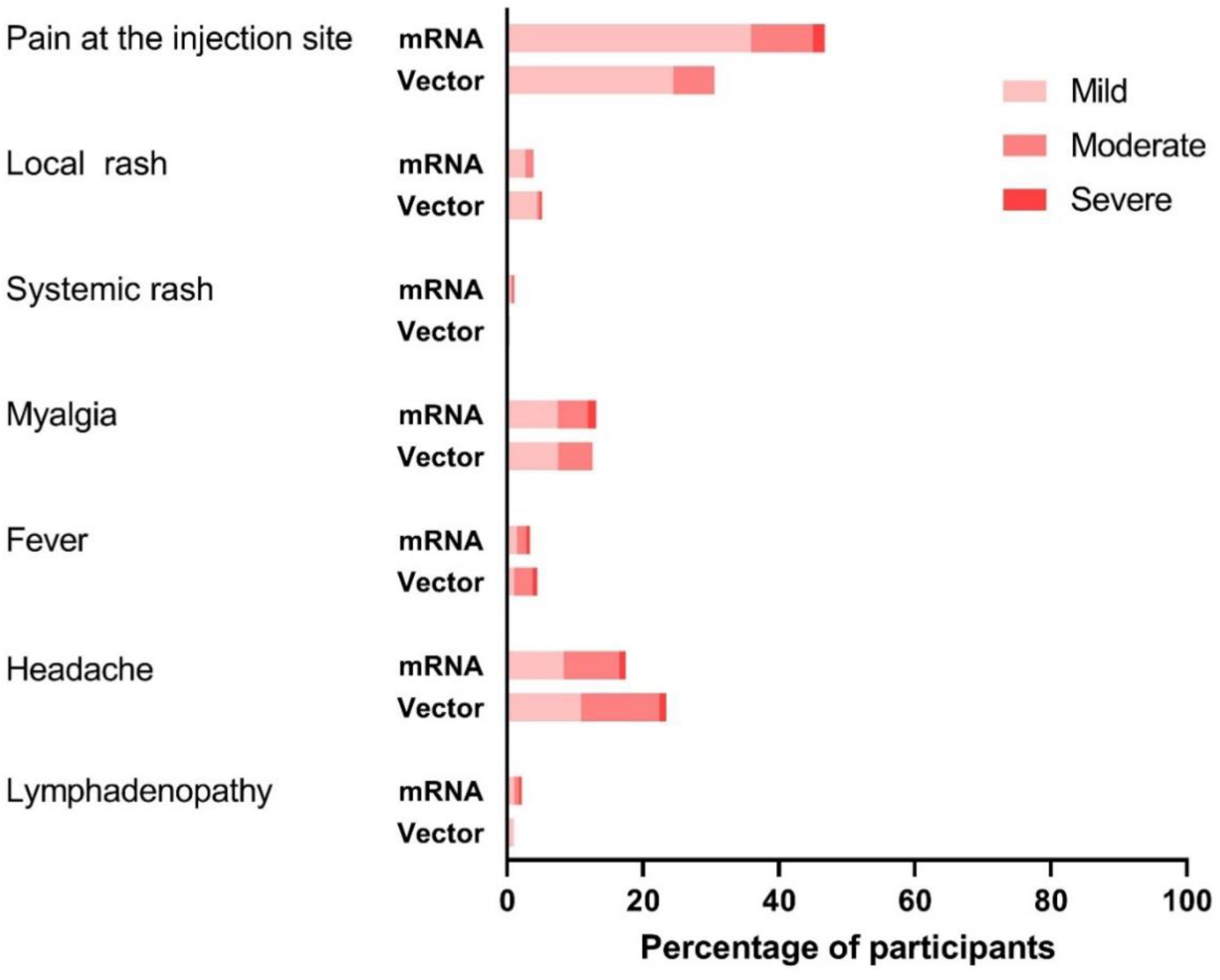
Severity of adverse events. Severity of AE present in the seven days after vaccination in PLWH comparing mRNA (BNT162b2, mRNA-1273) and vector (ChAdOx1-S, Ad26.COV2.S) vaccines shown as a percentage (%). AE: adverse events, PLWH: people living with HIV

## Conclusion

Limited data exist on SARS-CoV-2 vaccination responses in PLWH. Here, we show that mRNA induces lower SARS-CoV-2 S1-specific IgG levels in PLWH compared to controls, even after correction for age, sex, and vaccine type. PLWH receiving a vector vaccine, of an older age, and with lower CD4+ T-cell counts have more impaired antibody responses. As expected, the SARS-CoV-2 vaccines were well tolerated in PLWH without vaccine related discontinuations or SAE.

Our primary result analysis in participants receiving mRNA vaccines contrasts with most of the small cohort studies performed in PLWH where the authors found similar responses as in controls [16-18]. With regard to vector vaccines in our study, similar effects were seen as for mRNA vaccines, with lower antibody concentrations in PLWH compared to controls. This differs from previously published results on ChAdOx1-S where no differences were seen between PLWH and HIV-negative participants [14, 15]. Most, if not all, of the previous studies had not been powered to detect a predefined size of the impact of HIV on vaccination response. Therefore, this discrepancy is probably a type two error of the previous much smaller studies. Other reasons that may explain why some of the previous studies did not find a lower response in PLWH may be the use of qualitative rather than quantitative antibody responses and the inclusion of younger and more female participants. In contrast to the phase three trials, we observed a lower response in male participants [1, 2]. However, this was observed in other vaccination studies in PLWH previously [23]. Additionally, age is also known to influence the immune response to SARS-CoV-2. Our results confirm this in PLWH, in whom more immunosenescence is seen compared to HIV-negative controls [24, 25].

Overall, we found an increase in T-cell responses both on activation of CD4+ and CD8+ T-cells, and cytokine production when exposed to SARS-CoV-2 spike protein. There were still a relevant proportion of PLWH in whom T-cell responses after stimulation could not be measured. However, in the AIM assay this was also observed in controls. Low-response or non-response has also been seen in ELISPOT assays performed previously on healthy participants after vaccination [26]. We observed a relevant IFN-γ production in the negative control stimulation in the PLWH group. Chronic immune activation and persistent inflammation has been reported before in chronic HIV patients, and could explain these results [27].

Adverse events occurred in just over half of all cases. When looking at BNT162b2, the overall incidence of AE did not increase after the second vaccination although the type of AE differed somewhat (e.g. systemic events occurred more often after the second). Overall, AE patterns in PLWH were mild and similar to phase three trials, both in type and frequency [1-3, 5].

This study was performed at 22 of total 24 HIV treatment sites in the Netherlands. Our recruitment strategy resulted in a large group of PLWH, with reasonable representation of female, as well as elderly and those with lower CD4+ T-cell count. This reflected the HIV demographic in the Netherlands, in which the 90-90-90 goals were already reached in 2018 [28].

Several limitations are noteworthy. Because provision of vaccinations was decided by the Dutch National Institute for Public Health and the Environment, we could not fully control the distribution of the available vaccines across age, sex, and CD4+ T-cell strata. Furthermore, there were some differences in age and sex between the PLWH and controls, which we corrected for in our multivariate analyses. We can also not fully guarantee that all participants with an antecedent COVID-19 infection were excluded as antibodies may become undetectable over time. Finally, few PLWH with a very low CD4+ T-cell count were enrolled and even fewer with a viral load >50 copies/ml.

The COVID-19 landscape continues to change rapidly as new VOC are emerging. Recent studies have shown immune escape of the Omicron variant from humoral immunity induced by infection as well as vaccination [26]. However, at least in HIV-negative people, this can be overruled by a booster vaccination [21]. Additionally, higher antibody levels are associated with greater protection against symptomatic disease [29, 30]. This highlights the importance that additional vaccinations can play in controlling the pandemic. Based on the results of this study, we decided to give all participating PLWH with an antibody response below 300 BAU/ml the opportunity to receive an additional mRNA-1273 vaccination. Furthermore, given the safety of the mRNA vaccines, the overall lower vaccine inducible antibody response observed in PLWH, the observed waning of serum antibody levels over time, and immune escape by VOC, we think that providing additional vaccinations to all PLWH may optimise protection. An argument can be made for a more targeted approach in for example older PLWH, those with lower CD4+ T-cells, or based on measured antibody responses and neutralisation capacity after vaccination, however, we consider this too impractical.

In conclusion, vaccination against SARS-CoV-2 of PLWH resulted in a lower antibody response compared to HIV-negative controls. Additional vaccinations may therefore be required in order to compensate for this reduced antibody response.

## Data Availability

Individual participant data that underlie the results reported in this article, after de-identification, will be made available to researchers who provide a methodologically sound study proposal.

## Acknowledgements

Foremost, we would like to thank all participants of the study to help advancing science. We also want to thank the following people who helped in the recruitment of participants: Aniek Adams, A. Boonstra, Marjolein van Broekhuizen, Margo van der Burg-van der Plas, A. Cents-Bosma, Willemien Dorama, T. Duijf, S. Faber, Natasja van Holten, Astrid van Hulzen, L.M. Kampschreur, Annemarie van der Kraan, Inge de Kroon, Laura Laan, Eliane Leyten, Vera Maas, P.A. der Meulen, Femke Mollema, Suzanne de Munnik, Hans-Erik Nobel, Vincent Peters, Simone Phaf, M. Pietersma, Frank Pijnappel, Leontine M.M. van der Prijt, Linda Scheiberlich, Jasmijn Steiner, Jolanda M. van der Swaluw, Maartje Wagemaker, Annouschka Weijsenveld, Marc van Wijk, Sieds Wildenbeest and Sabine van Winden. We would like to thank all our colleague internist-infectious diseases specialists in the Netherlands who helped with the patient recruitment. Finally, we would like to thank Alessandro Sette and Alba Grifoni (La Jolla Institute for Immunology, La Jolla, San Diego, USA) for providing the peptide pools used in the AIM assay.

## Supporting Information

### S1 Appendix

#### Study Design and Participants

Inclusion was stratified according to vaccine type (mRNA, vector), sex assigned at birth, age (18-55, 56-65 and >65), and most recent CD4+ T-cell count (<350 and ≥350 cells/µL). In order to recruit a study population that best represents the Dutch population of PLWH, we continuously monitored recruitment across these strata and strata were closed for enrolment when a sufficient number had been recruited [1].

All adult inhabitants of the Netherlands received an invitation for voluntary first round of vaccination with either BNT162b2, mRNA-1273, ChAdOx1-S (AZD1222), or Ad26.COV2.S between January and August 2021. The type of vaccination offered in the Netherlands was decided by the National Institute for Public Health and the Environment and was dependent on vaccine availability, employment in healthcare, age, and comorbidities. The government started vaccinating all healthcare workers and people above 65 years old with BNT126b2 from the end of January 2021. Subsequently, from the middle of February, all people between 60 and 64 years were invited for vaccination with ChAdOx1-S. From March onwards, all people with a medical indication, which included PLWH, between 18 and 60 years old were invited for vaccination with either BNT162b2, ChAdOx1-S or mRNA-1273. However, on the 8th of April the Governmental Dutch Health council suspended vaccination with ChAdOx1-S in people under the age of 60 due to the possibility of blood clots with low levels of platelets. The interval between the BNT162b2 vaccines was six weeks and between the mRNA-1273 vaccines five weeks. Up to 20 May 2021 the interval between the ChAdOx1-S vaccines was 12 weeks, after which the interval was changed to between four and 12 weeks. The Ad26.COV2.S vaccine was administered only once.

Vaccination response data from HIV-negative controls were obtained from two separate concurrent studies. The first cohort consisted of health care workers from the Erasmus Medical Centre in

Rotterdam who were enrolled in a prospective cohort study (n=385). Health care workers received BNT162b2, mRNA-1273, ChAdOx1-S, or Ad26.COV2.S according to national regulations as described above. The second group consisted of participants who served as non-immunocompromised controls in the Vaccination Against COVID-19 in Primary Immune Deficiencies (VACOPID) study investigating the mRNA-1273 vaccine in people with inborne errors of immunity (IEI) (n=55). They received two mRNA-1273 vaccines four weeks apart with blood sampling four weeks after the second vaccination. None of the controls had a history of COVID-19.

#### Clinical Procedures

Study variables that were collected included year of birth, sex assigned at birth, current use of combination antiretroviral therapy (cART), most recent plasma HIV-RNA (copies/mL), most recent CD4+ T-cell count (cells/µL), and nadir CD4+ T-cell count (cells/ µL).

#### Laboratory Procedures

Serum samples before vaccination were analysed at the laboratory of the individual treating centres using either Wantai SARS-CoV-2 total Ig and IgM ELISAs (Beijing Wantai Biological Pharmacy Enterprise Co., Ltd. China), Abbott ARCHITECT SARS-CoV-2 IgG (Abbott Park, Illinois, U.S.A.), Siemens Atellica-IM IgG (sCOVG) SARS-CoV-2 Serology Assay (Siemens Healthineers Nederland B.V.), or Liaison by DiaSorin (Saluggia, Italy), depending on local availability and according to the manufacturer’s instructions.

ELISpot assays were performed on cryopreserved PBMCs using a commercial double colour kit (Cellular Technology Limited). ImmunoSpot plates were coated with capture antibodies and kept overnight at 4°C. The plates were washed one of the following overlapping peptide pools were added as stimuli to the appropriate wells, with culture media (CTL-Test® medium with 1% L-Glutamine): spike glycoprotein (S, PM-WCPV-S1), nucleocapsid (N) protein (PM-WCPV-NCAP-1), myelin-oligodendrocyte glycoprotein (MOG) (PM-MOG, negative control), and CEFX (PM-CEFX-2, positive control) (all from JPT) at a concentration of 2 µg/ml. PBMCs were plated at 200,000 cells per well and incubated 72 hours (9% CO2). Cells were removed, detection antibodies and enzymes for visualisation of the cytokines were added, after which plates were air dried in the dark prior to counting with the ImmunoSpot Image Analyzer.

Activation induced marker (AIM) assays were also performed on cryopreserved PBMCs. 1×10^6^ PBMCs were incubated with SARS-CoV-2 peptide pools (1µg/mL) and incubated at 37°C for 20 hours. SARS-CoV-2 peptide pools consist of 15-mers with 10 amino acid overlaps covering the entire S-protein of the WuhanHu1 (wild-type) (in PLWH) and D614G (wild-type) (in HIV-negative controls) or B.1.617.2 (delta) variant. PBMCs were stimulated with an equimolar amount DMSO as negative control or a combination of PMA (50 µg/mL) and Ionomycin (500 µg/mL) as positive control. Following stimulation, cells were stained and measured by flow cytometry (FACSLyric, BD). PBMC were stained at 4°C for 15 minutes with the following antibodies and dilutions: anti-CD3^PerCP^ (Clone SK7, BD, 1:25), anti-CD4^V50^ (Clone L200, BD, 1:50), anti-CD8^FITC^ (Clone DK25, Dako, 1:25), anti-CD45RA^PE-Cy7^ (Clone L48, BD, 1:50), anti-CCR7^BV711^, anti-CD69^APC-H7^ (Clone FN50, BD, 1:50), anti-CD137^PE^ (Clone 4B4-1, Miltenyi, 1:50), and anti-OX40^BV605^ (Clone L106, BD, 1:25). LIVE/DEAD™ Fixable Aqua Dead Cell staining was included (AmCyan, Invitrogen, 1:100).

#### Outcomes

Analysed variables associated with the height of antibody level; vaccine type (BNT162b2, mRNA-1273, ChAdOx1-S, or Ad26.COV2.S), sex assigned at birth (male or female), age (birth year subgroups 1965-2002, 1964-1955 and <1954), nadir CD4+ T-cell count (<250, 250-500 and >500 cells/µL), and most recent CD4+ T-cell count (<250, 250-500 and >500 cells/µL).

Analysed variables associated with hyporesponse and non-response; vaccine group (mRNA or vector), sex at birth (male or female), age group (birth year subgroups 1965-2002, 1964-1955 and <1954), nadir CD4+ T-cell count (<250, 250-500 and >500 cells/µL), and most recent CD4+ T-cell count (<250, 250-500 and >500 cells/µL).

We evaluated the tolerability of the administered vaccines by monitoring local and systemic vaccine related adverse events. Severity of reactogenicity was measured as mild (symptoms present but no functional impairment or medication needed), moderate (necessitating medication, no functional impairment) or severe (impairing in daily functioning). Severe adverse events (SAE) were assessed for likeliness of association with vaccination by the participating site principal investigator and physician.

---

**S1 Table:**
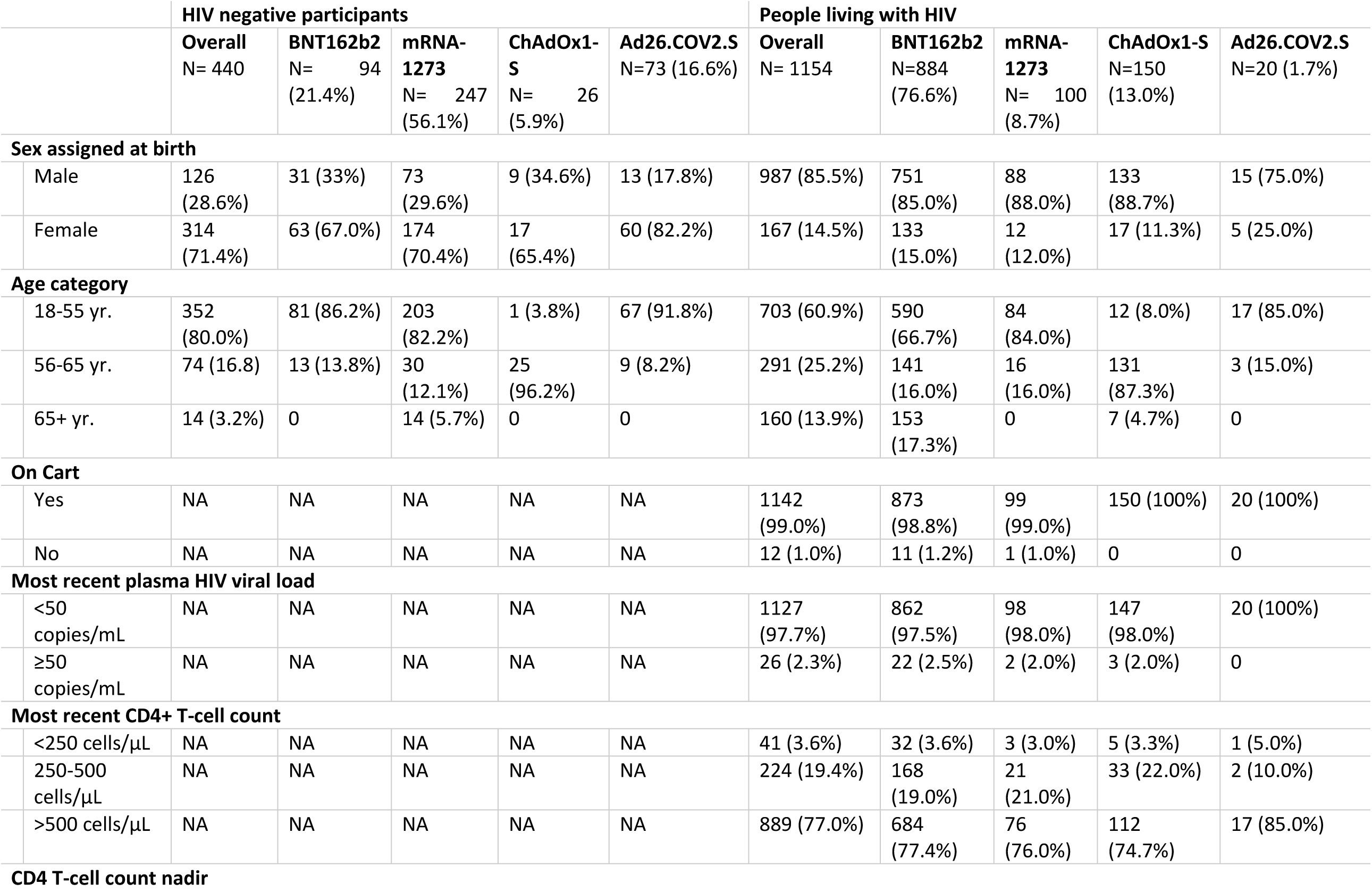
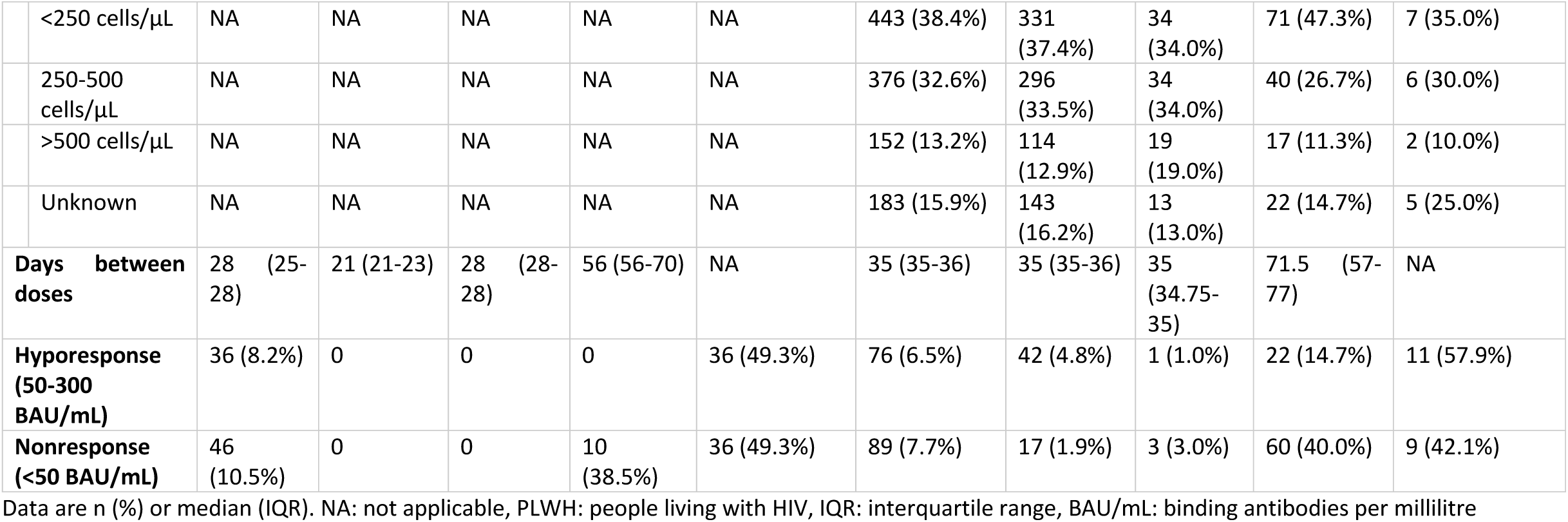
Baseline characteristics of HIV negative participants and PLWH per vaccine.

**S2 Table:**
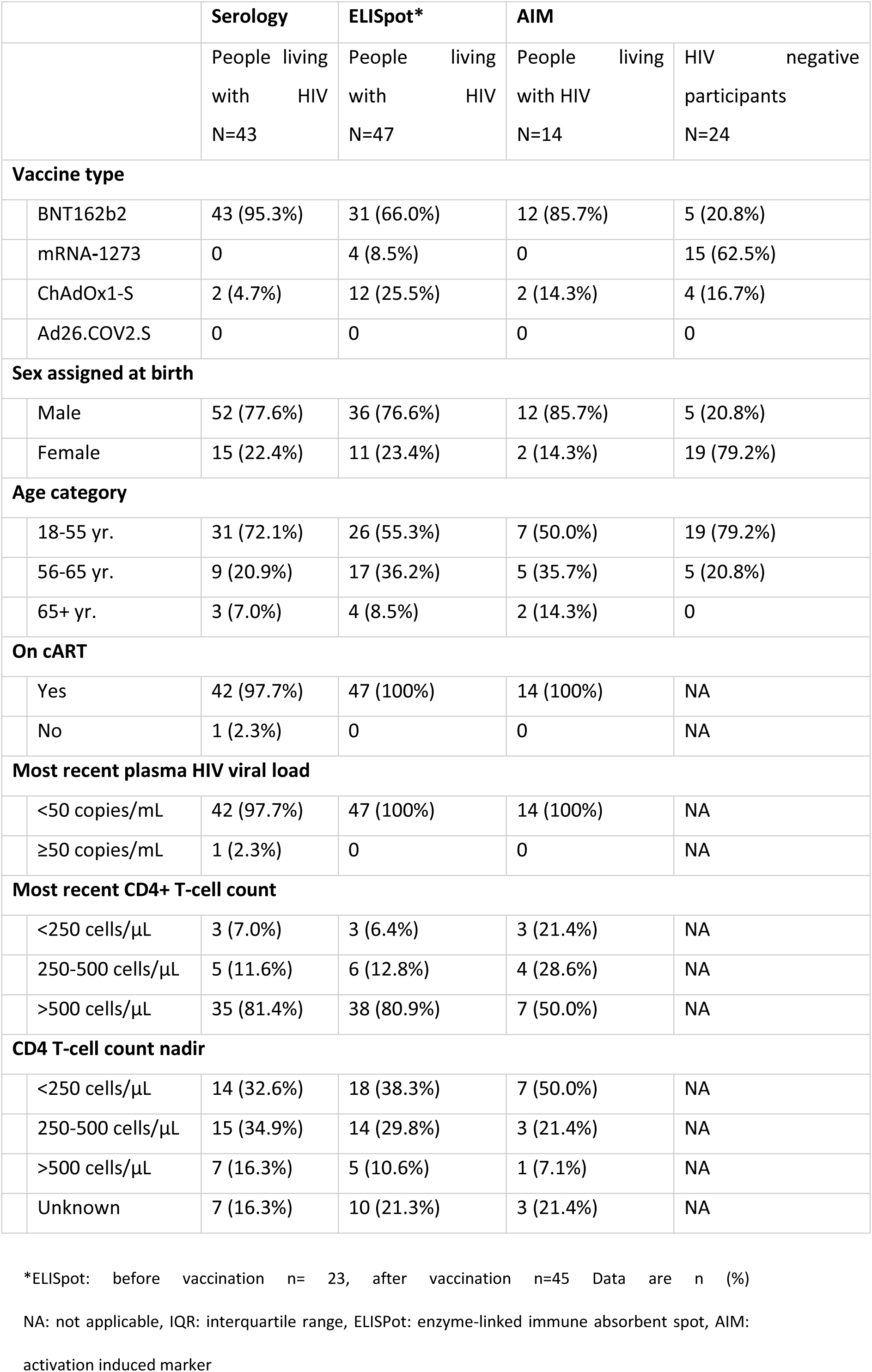
Subgroup patient characteristics.

**S1 Fig:**
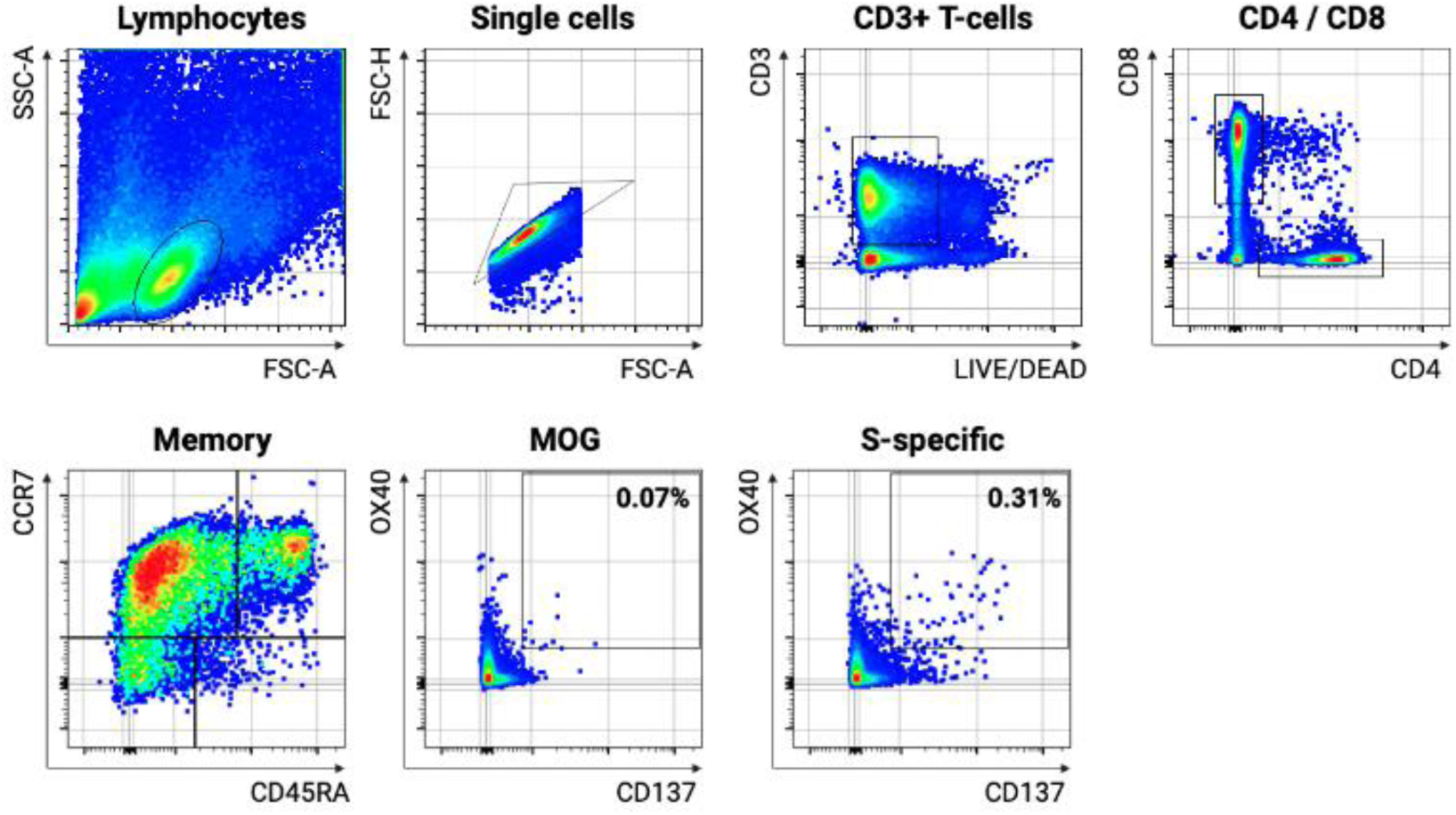
Gating strategy. Gating strategy for Flow Cytometry in AIM assay. AIM: activation induced marker, MOG: myelin-oligodendrocyte glycoprotein

**S2 Fig:**
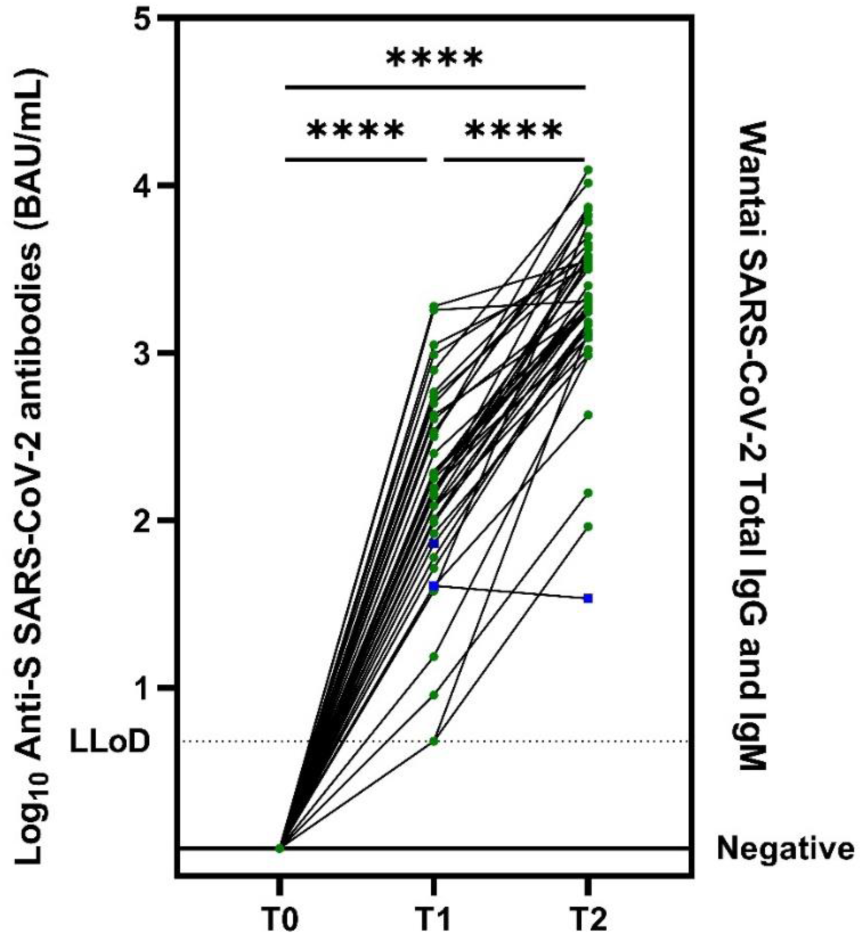
Serologic responses after vaccination in PLWH. T0: before vaccination, T1: 21 (+/-3) days after first vaccination, T2: 4-6 weeks after second vaccination Antibody concentration measured as S-specific binding antibodies per millilitre (BAU/ml) in the subgroup participants (n=43) (T0 is Wantai, all negative). Green circles: PLWH who received mRNA vaccines, Blue squares: PLWH who received vector based vaccines. The dotted line shows the lower limit of detection (4.81 BAU/ml), statistics performed with Wilcoxon matched-pairs signed rank test (**** p<0.0001). S: spike, BAU/ml: binding antibodies per millilitre, PLWH: people living with HIV, LLoD: lower limit of detection

**S3 Fig:**
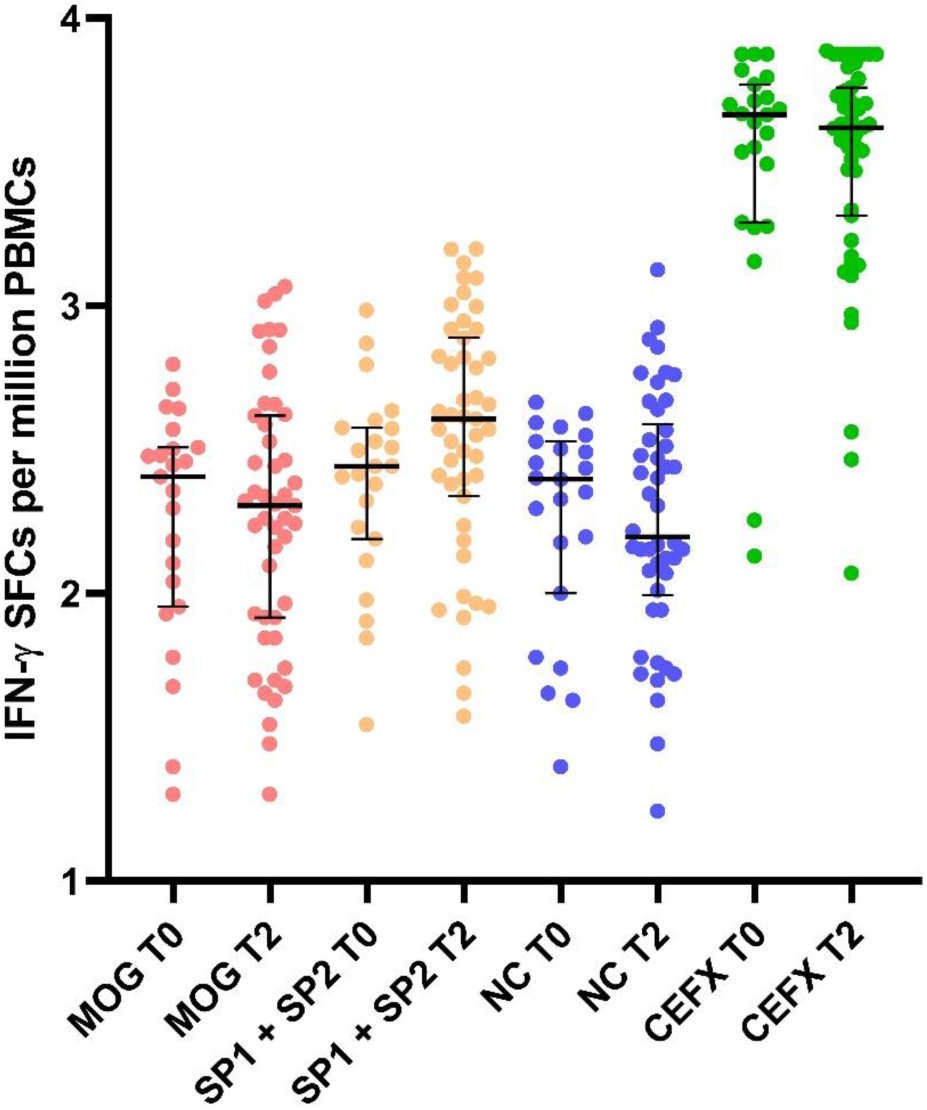
ELISPOT results in subgroup participants (PLWH) T0: before vaccination, T2: 4-6 weeks after second vaccination All ELISpot assay results shown as IFN-γ SFC. MOG: myelin-oligodendrocyte glycoprotein, SP1: spike protein 1, SP2: spike protein 2, NC: nucleocapsid, PLWH: people living with HIV, IFN: interferon, SFC: spot forming cells, PBMCs: peripheral blood mononuclear cells, ELISPot: enzyme-linked immune absorbent spot

